# Multi-ancestry Whole-exome Sequencing Study of Alcohol Use Disorder in Two Cohorts

**DOI:** 10.1101/2024.04.05.24305412

**Authors:** Lu Wang, Henry R. Kranzler, Joel Gelernter, Hang Zhou

**Affiliations:** Department of Psychiatry, Yale University School of Medicine, New Haven, CT; Veterans Affairs Connecticut Healthcare System, West Haven, CT; Department of Psychiatry, University of Pennsylvania Perelman School of Medicine, Philadelphia, PA; Crescenz Veterans Affairs Medical Center, Philadelphia, PA; Department of Genetics, Yale School of Medicine, New Haven, CT; Department of Neuroscience, Yale School of Medicine, New Haven, CT; Section of Biomedical Informatics and Data Science, Yale School of Medicine, New Haven, CT; Center for Brain and Mind Health, Yale School of Medicine, New Haven, CT

**Author notes:** <u>Corresponding Authors:</u> Hang Zhou, Department of Psychiatry, Yale School of Medicine; Veterans Affairs Connecticut Healthcare System Annex Center, room 1325, 200 Edison Rd, Orange, CT 06477, USA.; Joel Gelernter, Department of Psychiatry, Yale School of Medicine, Veterans Affairs Connecticut Healthcare System, 116A2, 950 Campbell Ave, West Haven, CT 06516, USA.

## Abstract

Alcohol use disorder (AUD) is a leading cause of death and disability worldwide. There has been substantial progress in identifying genetic variants underlying AUD. However, there are few whole-exome sequencing (WES) studies of AUD. We analyzed WES of 4,530 samples from the Yale-Penn cohort and 469,835 samples from the UK Biobank (UKB). After quality control, 1,420 AUD cases and 619 controls of European ancestry (EUR) and 1,142 cases and 608 controls of African ancestry (AFR) from Yale-Penn were retained for subsequent analyses. WES data from 415,617 EUR samples (12,861 cases), 6,142 AFR samples (130 cases) and 4,607 South Asian (SAS) samples (130 cases) from UKB were also analyzed. Single-variant association analysis identified the well-known functional variant rs1229984 in *ADH1B* (*P*=4.88×10^-31^) and several other common variants in *ADH1C*. Gene-based tests identified *ADH1B* (*P*=1.00×10^-31^), *ADH1C* (*P*=5.23×10^-7^), *CNST* (*P*=1.19×10^-6^), and *IFIT5* (3.74×10^-6^). This study extends our understanding of the genetic basis of AUD.

## Introduction

Heavy alcohol use and alcohol use disorder (AUD) are leading causes of death and disability worldwide. Globally, in 2020, alcohol use accounted for 1.78 million deaths^1^. AUD is a complex disorder affected by both environmental and genetic factors, with twin studies showing AUD to have an estimated heritability of ∼0.50^2^. Identifying genetic factors that contribute to AUD risk could advance efforts to prevent, identify, and treat the disorder and common co-occurring medical and psychiatric problems related to alcohol use.

Over the past 30 years, candidate gene studies of AUD have established the importance of the functional coding variants rs1229984 in *ADH1B* (alcohol dehydrogenase 1B (class I), beta polypeptide) in multiple populations, and rs671 in *ALDH2* (aldehyde dehydrogenase 2 family (mitochondrial))^3-6^ in some Asian populations. These two genes encode enzymes that play critical roles in ethanol metabolism^7^. There has also been substantial progress made in genetic studies of AUD and problematic alcohol use (PAU, a proxy phenotype of AUD) through genome-wide association studies (GWAS). Key findings have been obtained that were impossible before the big data era^8^. Dozens of risk variants have been discovered^9-15^, primarily in European-ancestry (EUR) subjects, which have improved causal inferences between related traits^15^. AUD differs from alcohol consumption genetically, to some extent each having distinct genetic correlations with other health-related traits^13,14^, supporting the particular medical significance of AUD. The partitioned heritability of AUD and gene expression is enriched in brain tissues, indicating that AUD involves brain pathology^14-16^.

We previously conducted a large PAU GWAS of more than one million individuals of multiple ancestries, which extended our understanding of the biology of AUD and improved potential translational applications^17^. In that study, we demonstrated a substantial shared genetic architecture of PAU across multiple ancestries therefore the cross-ancestry fine-mapping improved the identification of potential causal variants, and the cross-ancestry polygenic risk score (PRS) analysis showed better prediction than single-ancestry PRS. The study prioritized multiple genes with convergent evidence linking AUD risk to the brain. It also identified existing medications for potential pharmacological studies by drug-repurposing analysis.

Although these findings have considerably advanced our knowledge of the genetics of AUD, large gaps remain. The estimated single-nucleotide polymorphism (SNP)-based heritability (*h*^2^) of AUD ranges from 5.6% to 12.7% with liability-scale *h*^2^ ranging from 8.9% to 16.2%^12-15,17^, far less than estimates from genetic epidemiology. Increasingly, whole-exome sequencing (WES) data are being used to augment SNP data for many diseases and traits^18-22^. WES outperforms SNP-array-based studies in identifying rare variants and accounting for missing heritability^21,23-25^. Large-scale WES studies of patients with neuropsychiatric disorders such as bipolar disorder and schizophrenia have implicated ultra-rare variants (URVs) in coding regions as risk factors or found enrichments of protein-truncating variants from evolutionarily constrained genes^18,22^. However, the analysis of WES data in AUD has been limited to small samples because of the limited availability of AUD patients with genetic data^26,27^, thus there is a need for a large-scale WES study with greater power to detect risk variants for AUD. Thus, we performed the largest multi-ancestry WES study in AUD to date by combining data from the Yale-Penn cohort (YP)^11^ and UK Biobank (UKB)^28^. We confirmed previously observed associations of the coding variant rs1229984 with AUD and obtained suggestive evidence of several novel rare variants. Enrichment analyses applying Firth’s logistic regression showed a nominally significant burden of ultra-rare loss-of-function (LoF) variants in evolutionarily constrained genes (pLI>0.9) for AUD cases. Furthermore, using gene-based tests accounting for the burden from LoF, missense and synonymous variants, we identified the well-studied *ADH1B* and *ADH1C*, and novel genes *CNST* and *IFIT5*.

## Results

### WES datasets

After quality control (QC, see Methods), whole-exome sequencing (WES) data from 3,892 participants of the YP cohort was included, including 2,102 EUR individuals and 1,790 individuals of African ancestry (AFR) (Table 1). AUD diagnosis (DSM-IV^29^ alcohol dependence) was obtained using the Semi-Structured Assessment for Drug Dependence and Alcoholism (SSADDA)^30^. We analyzed 1,420 EUR cases and 619 EUR controls and 1,142 AFR cases and 608 AFR controls. SNPs and INDELs (insertions and deletions), were jointly called following a GATK Best Practices Workflow^31^, and a series of QC steps performed to remove low-quality samples and variants (Figure 1, Figure S1). Over 3 million variants passed QC, of which 13.22% are not reported either in the Exome Aggregation Consortium (ExAC)^32^ data or the Genome Aggregation Database (gnomAD) version 3.1.2^33^ (Figure S2). Among these novel variants, 1,988 were present only in EUR and 803 variants were specific to AFR. We found that 98.19% of the novel variants (154,592) were shared by EUR and AFR.

**Figure 1.**
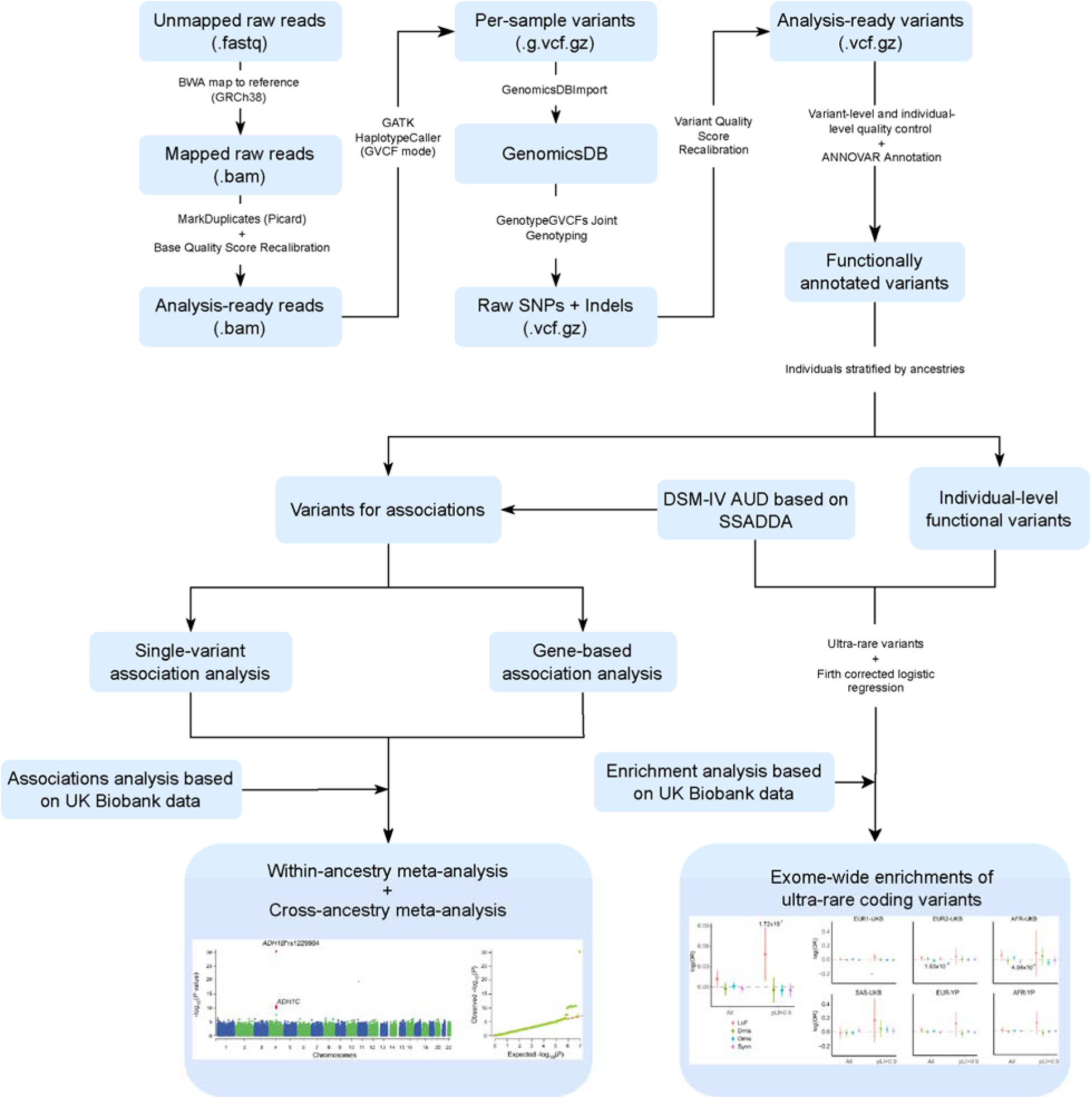
Workflow of this study. WES data from YP cohort went through the BWA-GATK variant calling pipeline and then association analyses were done at both single-variant and gene-based levels. Meta-analyses integrating UKB dataset have been performed for association results and enrichments of functional URVs across UKB and YP cohorts and ancestries. Logistic regression with Firth’s correction has been utilized to test the enrichments of each type of functional coding variants.

**Table 1.**
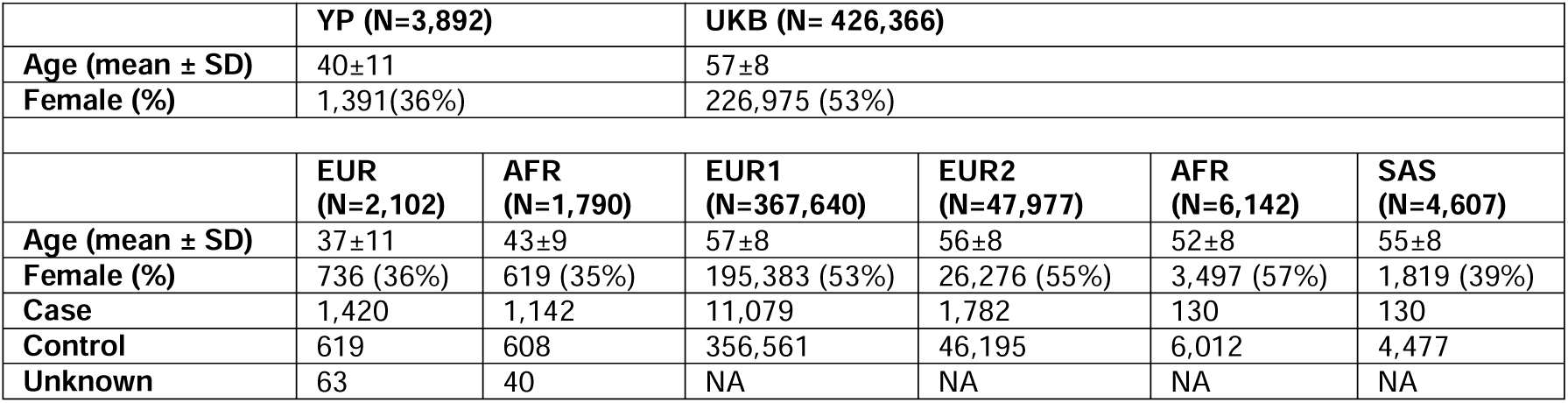
Demographic information for Yale-Penn and UK Biobank. Mean age ± standard deviation (SD) and proportions of females are shown for all participants from YP and UKB that have both phenotype and variant information. YP, Yale-Penn; UKB, UK Biobank; EUR, European; AFR, African; SAS, South Asian. NA, not applicable.

|  | YP (N=3,892) |  | UKB (N= 426,366) |  |  |  |
| --- | --- | --- | --- | --- | --- | --- |
| **Age (mean ± SD)** | 40±11 | | 57±8 | | | |
| **Female (%)** | 1,391(36%) |  | 226,975 (53%) |  |  |  |
|  | EUR<br>(N=2,102) | AFR<br>(N=1,790) | EUR1<br>(N=367,640) | EUR2<br>(N=47,977) | AFR<br>(N=6,142) | SAS<br>(N=4,607) |
| **Age (mean ± SD)** | 37±11 | 43±9 | 57±8 | 56±8 | 52±8 | 55±8 |
| **Female (%)** | 736 (36%) | 619 (35%) | 195,383 (53%) | 26,276 (55%) | 3,497 (57%) | 1,819 (39%) |
| **Case** | 1,420 | 1,142 | 11,079 | 1,782 | 130 | 130 |
| **Control** | 619 | 608 | 356,561 | 46,195 | 6,012 | 4,477 |
| **Unknown** | 63 | 40 | NA | NA | NA | NA |

WES data for 426,366 UK Biobank (UKB) participants were examined across EUR, AFR and South Asian (SAS) ancestries (Methods). UKB has assigned 367,640 British participants to EUR and hereafter are referred to as EUR1 subpopulation^28^. UKB participants without EUR1 ancestry were analyzed by principal component analysis (PCA), integrating subjects from the 1000 Genomes Project^34^ and 47,977 participants were then assigned to the an additional EUR group (hereafter referred as EUR2), 6,142 to AFR and 4,607 to SAS (details of this approach are described in our previous study^17^).

### Examining loss of function (LoF) variants in AUD

To ascertain whether deleterious coding variants were enriched in AUD cases or controls, we annotated the variants using ANNOVAR^35^ and categorized the variants according to these predictions into four groups based on the annotations: loss-of-function (LoF), damaging missense (Dmis), other missense (Omis) and synonymous (Synn) (Methods, Table S1). The deleteriousness of missense variants was determined by applying an ensemble pathogenicity predictor for exome missense variants (REVEL) with a cutoff of 0.5 to optimize sensitivity and specificity^36,37^. Logistic regressions with Firth’s correction^38^ were performed on URVs with minor allele count (MAC) ≤ 5 (Figure 2, Tables S2-3).

**Figure 2.**
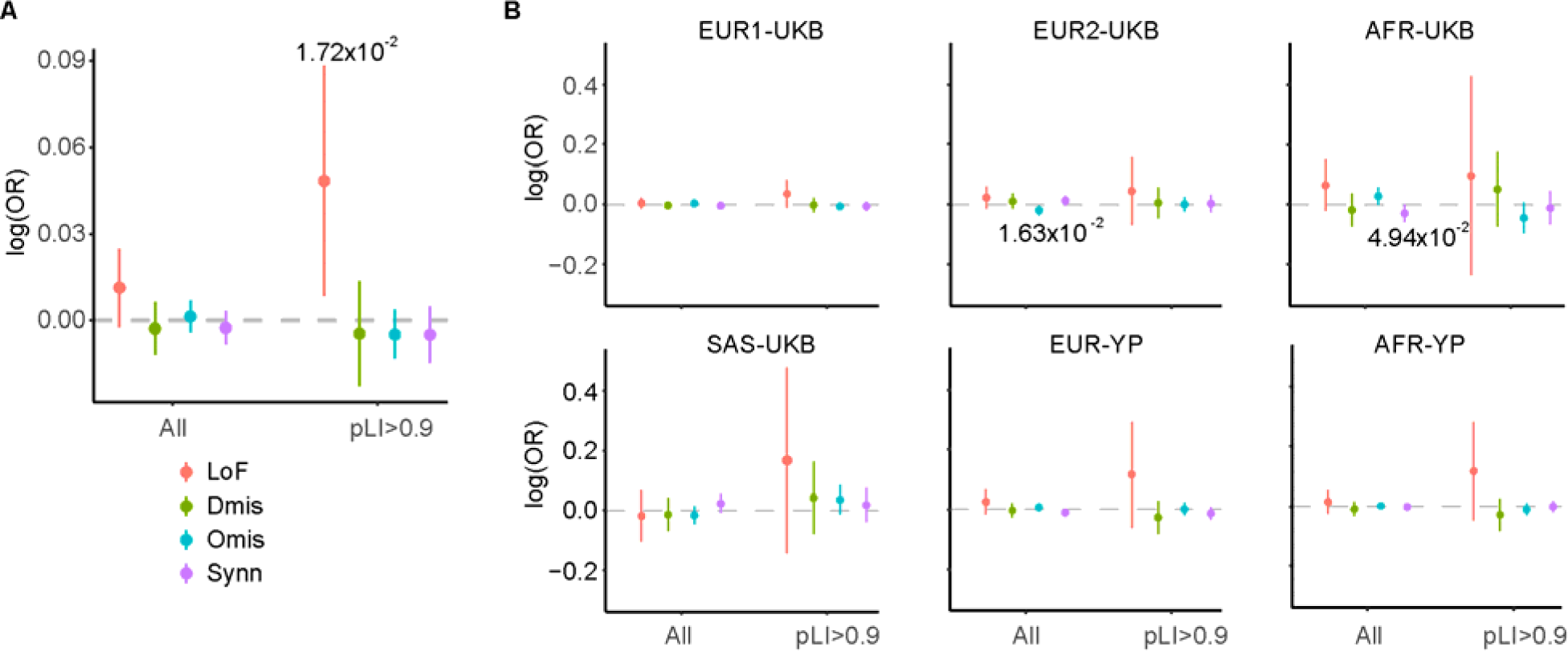
Functional enrichment analysis of URVs in YP and UKB. Case-control enrichment of URVs across functional categories of LoF, Dmis, Omis and Synn among all ancestry groups from YP and UKB. Enrichments were shown for all coding genes (N=21,162) and the evolutionarily constrained genes with pLI>0.9 (N=2,894). Odds ratios were calculated using logistic regression corrected with Firth’s method. Two-sided *P* values from Wald test were shown. Enrichment results were shown for combined dataset across YP and UKB (A), and for each subpopulation in YP and UKB separately (B).

Adjusted for batch, age, sex, 10 principal components and the overall coding variant burden, URVs were not significantly enriched for any class of variants in AUD or in controls in either EUR or AFR from the YP cohort (Figure 2B and Table S2). After meta-analyzing URV enrichments across all subpopulations from the UKB and YP cohorts, we observed nominally significant enrichment of LoF URVs in evolutionarily constrained (pLI > 0.9) genes (*P*=1.72×10^-2^, Figure 2A, Table S3).

### Single-variant association analysis

We used a logistic mixed model implemented in SAIGE-GENE+^39^ to assess the association of exome-wide variants with AUD (Table S4). Single-variant association analyses showed no significant signals in the YP cohort (Table S5-6). The well-established AUD-associated SNP rs1229984 in the *ADH1B* gene was ranked as the top signal in EUR (*P*=7.95×10^-7^; Figure S3) but did not pass the threshold after Bonferroni correction for exome-wide multiple testing (*P*_Bonferroni_ = 4.13×10^-7^). In UKB, rs1229984 was significantly associated with AUD risk in the largest sub-population, EUR1 (*P*=1.21×10^-27^, Figure S4). We did not observe any significant signals from EUR2, AFR or SAS in UKB by single-variant association analyses (Figure S4, Tables S7-10).

### Within- and cross-ancestry meta-analyses

We next performed within- and cross-ancestry meta-analyses combining YP and UKB. Of the 8,129,748 variants in EUR, rs1229984 in *ADH1B* gene (*P*=2.84×10^-29^) and 9 variants in *ADH1C* gene were significantly associated with AUD at a Bonferroni-corrected threshold of *P*=6.15×10^-9^, with 3 additional variants in *ADH1C* meeting the FDR-adjusted *P*-value threshold of 2.04×10^-8^ (Table S11). No significant variants were identified in the AFR meta-analysis (Table S12). The results from a cross-ancestry meta-analysis, mostly driven by information in EUR, showed significant signals consistent with what was observed in EUR-only, with *ADH1B\**rs1229984 identified as the lead SNP (*P*=4.88×10^-31^; Figure 3, Table S13). Using conditional analysis we examined the independence of the top variants that passed Bonferroni correction in the cross-ancestry meta-analysis (Table S14). Among these 10 variants, 3 were reported by both of two previous phenome-wide association studies based on exome-sequencing data from UKB.^20,40^ All these variants were reported in our previous PAU study (Table S14).^17^

**Figure 3.**
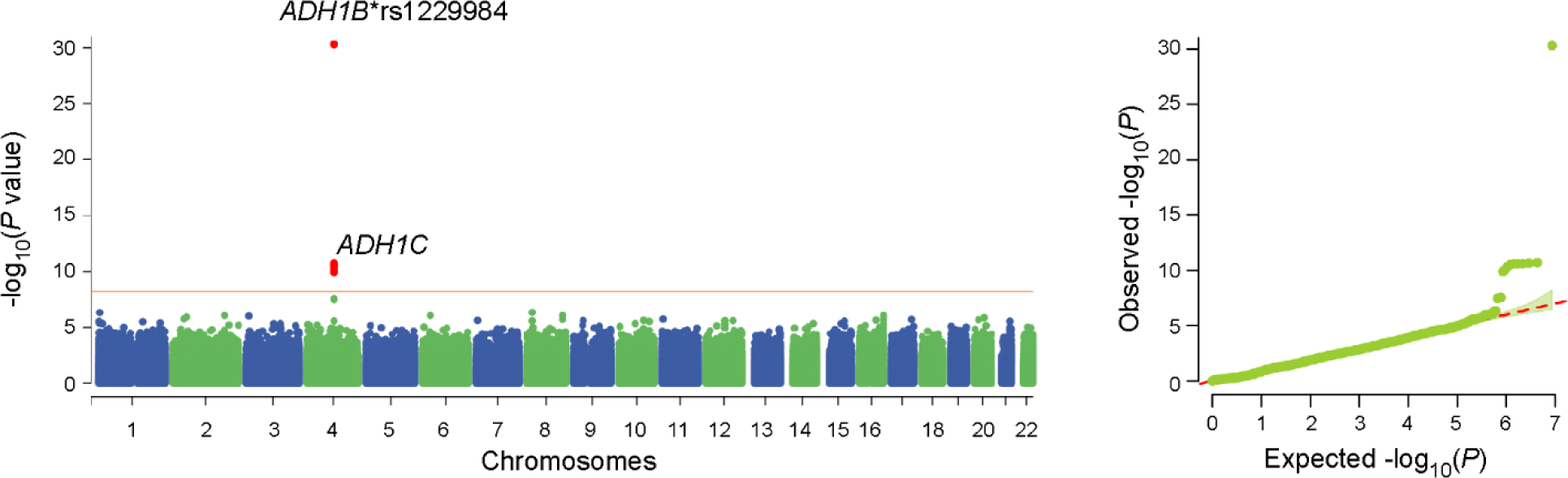
Manhattan and QQ plot for cross-ancestry meta-analyzed single-variant analysis across YP and UKB. A total of 8,590,046 variants across EUR1, EUR2, AFR and SAS from UKB and EUR, AFR from YP were analyzed. *N*_case_ = 13,121, *N*_control_ = 413,245.

### Gene-based analysis

Rare variants may act in aggregate and therefore could be detected by gene-based analysis rather than single-variant association analyses. Thus, we performed collapsing analyses to test the gene-level associations using variant aggregation strategies implemented in SAIGE-GENE+^39^ across multiple minor allele frequency (MAF) cutoffs and functional annotations. The predicted functional effects for variants were annotated using ANNOVAR^35^, as done in the enrichment analysis. For each gene, four separate variant burden masks were evaluated: LoF, “LoF|Missense”, “LoF|Missense|Synonymous”, and Synonymous (Table S1). For each of the masks, we considered different MAF cutoffs for each gene: MAF≤0.01%, MAF≤0.1%, MAF≤1% and all coding variants (in YP we restricted to MAF≤0.1%). The conservative Bonferroni-corrected exome-wide significance threshold was *P*≈2.36×10^-6^, corresponding to 21,162 genes tested in this study.

In the UKB EUR1 subpopulation, we observed exome-wide significant associations in *ADH1B* under the burden of all coding variants for “LoF|Missense|Synonymous” and “LoF|Missense” masks (*P*=3.70×10^-31^ and *P*=5.45×10^-31^, respectively; Figure S5). *CNST* (consortin, connexin sorting protein) was identified as close to the exome-wide gene-level significance under the burden mask of “LoF|Missense|Synonymous” for the rarest category of coding variants with MAF≤0.01% (*P*=2.75×10^-6^; Figure S5, Table S15). *OCA2* (oculocutaneous albinism II) was significantly associated with AUD in the AFR subpopulation for rare “LoF|Missense” variants with MAF≤1% (*P*=2.31×10^-6^; Figure S5, Table S16). No significant genes were identified in the UKB EUR2 and SAS subpopulations or the YP cohort (Tables S15-20).

Meta-analysis of EUR subjects across UKB and YP showed significant associations in *ADH1B* for “LoF|Missense” (*P*=2.66×10^-30^; Table 2, Table S21; Figure S6) and for “LoF|Missense|Synonymous” (*P*=6.02×10^-27^; Table 2, Table S21) under all coding variant masks. *ADH1C* showed significant associations for all coding LoF variants (*P*=1.99×10^-6^; Table 2, Table S21). We found no significant gene-level associations in the AFR meta-analysis, presumably due to limited statistical power (Table S22).

**Table 2.** Meta-analyses for gene-based tests at FDR-adjusted *P*-value < 0.05. LoF, loss-of-unction; MAF, minor allele frequency.

| Genes | Raw $P$ | FDR-adjusted $P$ | Category | MAF bin | Samples |
| --- | --- | --- | --- | --- | --- |
| *ADH1B* | $1.00 \times 10^{-31}$ | $2.05 \times 10^{-27}$ | LoF+Missense | 0.5 | Cross-ancestry |
| | $1.96 \times 10^{-30}$ | $4.01 \times 10^{-26}$ | LoF+Missense | 0.5 | Cross-ancestry UKB |
| | $2.66 \times 10^{-30}$ | $5.41 \times 10^{-26}$ | LoF+Missense | 0.5 | Within-ancestry EUR |
| | $6.02 \times 10^{-27}$ | $1.25 \times 10^{-22}$ | LoF+Missense+Synonymous | 0.5 | Within-ancestry EUR |
| | $1.22 \times 10^{-26}$ | $2.56 \times 10^{-22}$ | LoF+Missense+Synonymous | 0.5 | Cross-ancestry |
| | $6.66 \times 10^{-26}$ | $1.39 \times 10^{-21}$ | LoF+Missense+Synonymous | 0.5 | Cross-ancestry UKB |
| *ADH1C* | $5.23 \times 10^{-7}$ | $9.57 \times 10^{-3}$ | LoF | 0.5 | Cross-ancestry |
| | $8.87 \times 10^{-7}$ | $9.07 \times 10^{-3}$ | LoF+Missense | 0.5 | Cross-ancestry |
| | $1.99 \times 10^{-6}$ | $3.63 \times 10^{-2}$ | LoF | 0.5 | Within-ancestry EUR |
| | $2.54 \times 10^{-6}$ | $2.58 \times 10^{-2}$ | LoF+Missense | 0.5 | Within-ancestry EUR |
| | $3.04 \times 10^{-6}$ | $2.93 \times 10^{-2}$ | LoF+Missense | 0.5 | Cross-ancestry UKB |
| *IFIT5* | $3.74 \times 10^{-6}$ | $2.55 \times 10^{-2}$ | LoF+Missense | 0.5 | Cross-ancestry |
| | $4.31 \times 10^{-6}$ | $2.93 \times 10^{-2}$ | LoF+Missense | 0.5 | Cross-ancestry UKB |
| *CNST* | $1.19 \times 10^{-6}$ | $2.49 \times 10^{-2}$ | LoF+Missense+Synonymous | 0.001 | Cross-ancestry |
| | $2.09 \times 10^{-6}$ | $4.37 \times 10^{-2}$ | LoF+Missense+Synonymous | 0.001 | Cross-ancestry UKB |
| | $2.39 \times 10^{-6}$ | $4.97 \times 10^{-2}$ | LoF+Missense+Synonymous | 0.0001 | Cross-ancestry UKB |

Cross-ancestry meta-analysis of all UKB and YP subpopulations confirmed the associations of *ADH1B* and *ADH1C* with AUD (Figure 4 and Table 2). For rare variants with MAF≤0.1% under the “LoF|Missense|Synonymous” mask, the most significant gene-based association was identified in *CNST* (*P*=1.19×10^-6^, Table S23), for which significant associations have also been observed for the rarer variants under the MAF bin of 1×10^-4^ in UKB alone (Figures S5, Table S15). Under the gene burden mask of all LoF variants, *ADH1C* was most significantly associated with AUD (*P*=5.23×10^-7^, Table S23). *IFIT5* was significant at an FDR level < 5% for risk effects from all LoF and missense variants (*P*=3.74×10^-6^).

**Figure 4.**
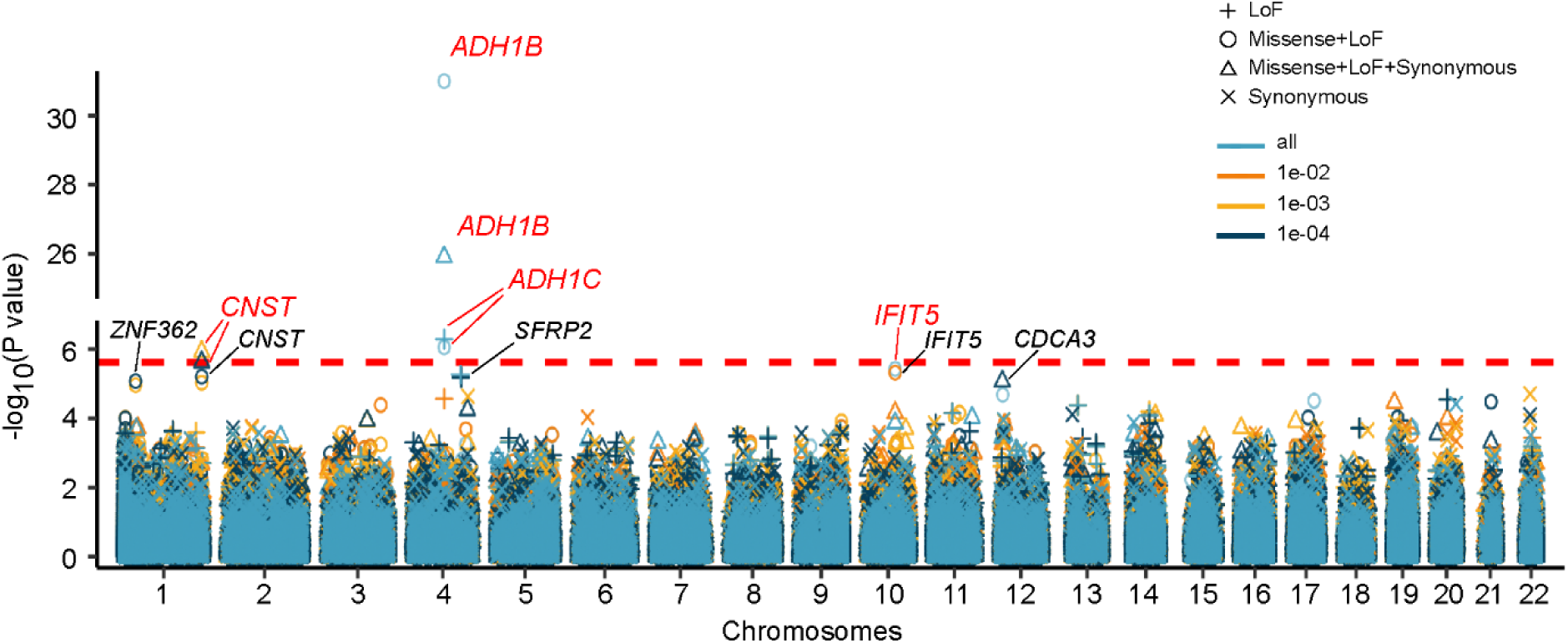
Meta-analyses of gene-based tests. Manhattan plot for meta-analyses of gene-based tests across different variant burden groups and MAF cutoffs. Red dashed line represented the Bonferroni-corrected significance threshold. Significant markers passing Bonferroni correction were highlighted in red. Markers with FDR-corrected *P*-value<0.1 were marked in black. Different shapes of dots indicated different variant groups while colors of dots represented MAF cutoffs.

Next, we extracted the genes that were nominally significant in at least one of the MAF and variant class combinations to perform Gene Ontology (GO) enrichment analysis. Although no individual GO term was significant after correction for multiple testing, “primary alcohol metabolic process” was the most enriched term of nominal significance (*P*=2.05×10^-4^; Figure S7).

### Sex-stratified analysis

We performed sex-stratified analysis across the UKB and YP cohorts for single-variant association tests (Tables S24-29). No significant signals were found for YP cohort males or females. In the UKB, *ADH1B*\*rs1229984 surpassed the Bonferroni corrected threshold only in males from the EUR1 subpopulation (*P*=5.97×10^-21^). A novel rare coding variant—rs756613425 in gene *U2AF1L4*—survived multiple testing correction (*P*=5.38×10^-9^) among females in EUR2. After meta-analysis, only rs1229984 in *ADH1B* remained significant in males in EUR populations (*P*=3.63×10^-22^, Table S24) and the cross-ancestry analysis (*P*=1.44×10^-23^, Table S28).

## Discussion

Rare variants play important roles in disease pathogenesis for some traits and characteristically, those that can be identified have larger effects than common variants^20,21^. Many causal variants for Mendelian diseases have been identified through rare variant analysis, e.g., *DHODH* (MIM: 126064) for postaxial acrofacial dysostosis (MIM: 263750)^41^. Rare coding variants detected by WES could explain additional disease risk or trait variability, beyond what is detected in common variant analyses. Studies have shown that rare variants or low-frequency variants (typically those with MAF<0.01) can explain roughly the same amount of variance in some traits as higher allele frequency SNPs^23,25^. A recent study using genome-based restricted maximum likelihood (GREML) on whole-genome sequence data from 21,620 subjects showed that SNPs with a MAF between 0.0001 and 0.001 explained more of the variance for height and BMI than many other allele frequency bins. Combining all common and rare variants fully recovered the heritability of height and BMI^23^. This may also be possible for other complex traits.

WES is a sequencing method that captures most protein-coding regions of the genome to identify exonic variants associated with a trait. An analysis of ∼450K UKB subjects showed that gene targets of drugs approved by the FDA were 3.6-fold more common among the associated genes by rare variant analysis than in the remaining genes, indicating the method’s potential clinical utility^21^. However, there is not much known about the AUD risk attributed to rare variants.

In this study, we present results from the largest multi-ancestry AUD WES study to date utilizing the YP and the UKB, which included 15,683 cases and 414,472 controls, facilitating our understanding of the genetic architecture of AUD. YP is a deeply phenotyped cohort with EUR and AFR participants recruited for genetic studies of substance use disorders^11^. The array-based genotype data from YP have contributed to several significant GWAS of AUD^12,17^. The YP WES data included in this study represent the largest purpose-recruited WES cohort for AUD. UKB is a population cohort with deep genomic and phenotyping data, comprising mostly EUR participants^28^. The prevalence of AUD in UKB is low compared to YP.

We confirmed the associations of the common coding variant *ADH1B\**rs1229984 and the genes *ADH1B* and *ADH1C* with AUD risk using both single-variant and gene-based association analyses. Inclusion of non-EUR ancestries enabled us to identify the novel gene *OCA2* in AFR, where the genetic burden identified by gene-based analysis fell on rare coding variants with MAF<0.01. Cross-ancestry meta-analysis of the gene-based results showed significant associations of another two novel genes, *CNST* (Consortin, Connexin Sorting Protein), conferred by coding variants with MAF<0.001, and *IFIT5* (interferon induced protein with tetratricopeptide repeats 5), conferred by LoF and missense variants. Sex-stratified analysis using UKB data identified a rare (MAF=1.14×10^-4^) protein-altering variant *U2AF1L4*\*rs756613425 in EUR2 females—neither the variant nor the gene has previously been associated with AUD (*P*=5.38×10^-9^, *P*_Bonferroni_=0.036).

In our cross-ancestry single-variant meta-analysis, suggestive (*P*<1×10^-6^) rare variants have been identified including rs117666725 (*SLC18A1*), rs1383727003 (*ARHGEF16*), and rs1437880 (*ASNSD1*). A previous association study has implicated *SLC18A1* with alcohol withdrawal severity but is attributable to a common SNP^42^. *ARHGEF16* is a guanine-exchange factor belonging to the Rho family GTPases, which plays important roles in neuron development and neurodegeneration^43^. *ASNSD1* encodes asparagine synthetase domain containing 1, which has been implicated in Huntington’s disease, a dominantly inherited neurological disorder^44^. None of these variants or genes have been identified to be associated with AUD risks in previous studies.

*IFIT5* has been reported to be involved in NF-ĸB pathway activation and promoting epithelial-to-mesenchymal transition via miRNA processing^45,46^. Previous studies on opioid use disorder and neuropathic pain implicated *IFIT5* with network analysis^47,48^. *CNST*, the consortin gene encoding the connexin sorting protein, was recently identified to be differentially expressed in anorexia nervosa, a serious mental disorder characterized by a high risk of mortality and morbidity^49,50^. Our gene-based test in AFR-UKB identified *OCA2* gene in AUD cases. *OCA2* is not just a primary pigmentation gene influencing the amount and characteristics of melanin in melanocytes, but it also stands as an independent factor impacting eye color^51^. Studies have shown that *OCA2* could be related to both eye color and neurodevelopmental disorders^52-54^. These genes could be novel targets for further functional studies for AUD.

Several limitations of our study should be noted. The enrichment analysis for different functional classes of URVs across ancestries only yielded nominal significant findings in LoF variants from evolutionarily constraint genes (Figure 2). This is considered to be most likely due to inadequate statistical power to identify the effects of deleterious URVs. Larger sample resources especially non-European genomic resources are urgently needed not only to delineate the contributions to AUD risk arising from deleterious URVs in coding regions, but also to more explicitly examine and compare the genetic variations across populations. Furthermore, WES only captures coding regions, which represent less than 2% of the human genome. To comprehensively and systematically investigate the particular genetic factors and their regulatory mechanisms underlying AUD risks, whole-genome sequencing offers an attractive approach. In addition, our analysis did not consider potential confounding factors such as socioeconomic status.

Our large-scale WES study is an advance in GWAS approaches to AUD that confirmed the associations of established variants with AUD and identified novel variants and genes by analyzing multiple ancestral groups. Future integrated studies of larger samples across multiple ancestries can be expected to advance our understanding of the multifaceted pathogenesis of AUD.

## Methods

### Ethics

The Yale–Penn study was approved by Yale Human Research Protection Program and University of Pennsylvania IRB.

### YP cohort

Whole-exome sequencing data for YP batch 1 cohort was generated on the Illumina HiSeq 2000 platform platform (paired end, 75 bp) with NimbleGen SeqCap exome capture V2 kit. The batch 2 to batch 4 data was sequenced on the NovaSeq 100bp paired end sequencing system using the xGen Exome Research Panel v1 from IDT. AUD cases were defined as participants with Diagnostic and Statistical Manual of Mental Disorders, Fourth Edition (DSM-IV) alcohol dependence diagnosed using the Semi-structured Assessment for Drug Dependence and Alcoholism (SSADDA).^30^ Principal component analysis was applied to the ancestry assignment of YP cohort integrating 1000 Genomes phase 3 subjects^34^ based on autosomal variants that remained after LD pruning.

Variants from the whole-exome sequences in the YP cohort were called following the BWA-GATK pipeline. Sequences were mapped to hg38 reference genome using BWA^55^. GATK HaplotypeCaller with GVCF mode was utilized to call individual variants. Next, GATK GenomicsDBImport and GenotypeGVCFs were applied to jointly genotype all samples, followed by VariantRecalibrator and ApplyVQSR to recalibrate variant quality scores. Variants located in sites with more than 6 alleles and low-complexity regions were filtered out. Heterozygous variants were removed if the sum of reference and alternative allele depth out of total sequencing depth of that site was below 0.8, the proportion of alternative allele depth was below 0.2, the Phred quality score of the reference allele was lower than 20, or the sequencing depth of the site was<10. For homozygous sites, variants with Phred quality score<20 or sequencing depth<10 were removed. Homozygous alternative allele sites with the proportion of sequencing depth for alternative alleles<0.8 were also removed. We used PLINK to remove variants with missingness rates>0.05; remove samples with inconsistent sex, highly related samples, or heterozygosity rate outside the mean ± 3 standard deviation range; and to filter out variants that failed Hardy-Weinberg equilibrium expectations (*P*<10^-6^). Samples with a missingness rate>10%, with mean depth<20, or with mean genotype quality score <55 were not retained for analysis. This resulted in the exclusion of 148 samples (Figure S1).

### UKB dataset

UKB is a large-scale prospective biomedical dataset that includes about 500,000 participants recruited throughout the United Kingdom. Exome-sequencing data from 469,835 participants were accessed. EUR1 refers to the genetically defined white British ancestry samples from UKB. Principal component analysis (PCA) integrating 1000 Genomes subjects was applied to define the EUR non-White-British (EUR2), AFR and SAS ancestries, as in our previous study^17^. The phenotype definition for UKB was based on data field 41270 (the ICD-10 diagnosis codes), and 20406 (self-report of ever having been addicted to alcohol). 10,466 participants had at least one F10 ICD-10 code, and an additional 3,593 participants self-reported to be addicted to alcohol were also defined as AUD cases. Participants reported to be lifetime abstainers or former drinkers (data field 20117), which might bias the genetic results as we demonstrated previously^56^, were removed from the control group. Participants reported to be current drinkers (data field 20117) were included as control samples. The final UKB analysis dataset comprised 426,366 samples across four subpopulations (367,640 EUR1, 47,977 EUR2, 6,142 AFR and 4,607 SAS). Variants with a missingness rate<5% were retained for subsequent analysis. PLINK was used to exclude variants that failed the Hardy-Weinberg equilibrium expectations with *P* <1×10^-20^, which was assessed separately in cases and controls.

### Variant annotation

We applied ANNOVAR to annotate all the variants passing the quality control steps according to predicted functional effect with RefSeq genome assembly hg38 release, cytoBand and avsnp150 database^35^. Variants predicted to be frameshift substitution, stop gain, stop loss or splicing site alterations were categorized into LoF variants. The value of 0.5 evaluated by REVEL^36^ was used as a cutoff for determining deleteriousness of missense variants. Variants with predicted REVEL score over 0.5 were considered as deleterious missense (Dmis) variants. Other missense (Omis) variants were those with REVEL score ≤0.5.

### Single-variant association analysis

SAIGE-GENE+ was used to perform single-variant association analysis, restricted to variants with MAC ≥ 5. For UKB, data analysis, sex, age and 10 ancestry principal components (PCs) were used as covariates. For the YP cohort, covariates included for analysis were age, sex, 10 PCs and sequencing batch information. Single-variant association analyses were performed for EUR and AFR for YP cohort, and EUR1, EUR2, AFR, SAS subpopulations for UKB. *P*-values corrected by the Bonferroni method were used as the genome-wide significance threshold for the independent tests from each single-variant association analysis to prevent possible false positive signals. FDR-adjusted *P*-values were also calculated to examine whether there were any variants that satisfied a less conservative threshold (See Tables S5-13).

### Conditional analysis

We evaluated the independence of significant variants by performing conditional analysis using SAIGE-GENE+^39^ on chromosome 4 in the EUR1 subpopulation from UKB. Ten candidate variants that passed the Bonferroni correction were tested (Table S14). The files used as input to store the null model and variance ratios derived from genetic relationship matrices based upon multiple minor allele count categories for running SAIGE-GENE+ were the same as those calculated in previous single-variant analyses.

### Gene-based tests

Rare coding variants were aggregated by different burden masks (LoF, “LoF| Missense”, “LoF|Missense|Synonymous”, Synonymous) for each gene. We tested the four gene masks across different allele frequency groups (MAF≤0.01%, MAF≤0.1%, MAF≤1% and all) using SAIGE-GENE+^39^. Covariates utilized for regression analysis were the same as those used in single-variant analysis. The Bonferroni-corrected significance threshold for exome-wide gene-based analysis was set at *P*≈2.36×10^-6^ (0.05 divided by a total of 21,162 genes from those included in the analyses of YP and UKB).

### Enrichment of functional ultra-rare variants (URVs)

URVs were defined as those with minor allele count (MAC) ≤ 5 in the corresponding dataset. Evolutionarily constrained genes were defined as those intolerant to LoF variants with pLI (the probability of being LoF intolerant) ≥ 0.9. Logistic regression with Firth’s bias reduction procedure was applied to compare enrichment of URVs and URVs in evolutionarily constrained genes among AUD cases and controls, adjusted for age, sex, 10 PCs in UKB and age, sex, 10 PCs and batch information for YP. Baseline variants that represented the total number of analyzed coding variants (LoF, missense and synonymous) were also incorporated as a covariate. The combined enrichment results for each functional class of variants across the subpopulations from UKB and YP cohort were evaluated using an inverse-variance weighted meta-analysis implemented in METAL^57^.

### Meta-analyses

Within-ancestry meta-analyses for EUR and AFR and a cross-ancestry meta-analysis of single-variant associations and gene-based tests were conducted using METAL^57^ with a sample-sized weighted approach. Meta-analyzed gene-based results were performed for each gene burden mask group across different allele frequencies. For MAF<0.01% variants across each burden mask group, cross-ancestry meta-analysis was performed only in UKB. For MAF<0.1%, MAF<1% variants and all variants, within-ancestry meta-analyses in European or African ancestry and cross-ancestry analyses were performed across UKB and YP.

## Data Availability

All data produced in the present study are available upon reasonable request to the authors

## Acknowledgments

This research used data from the UK Biobank (project ID: 41910), a population-based sample of participants whose contributions we gratefully acknowledge. The data access is supported by Yale-SCORE on sex differences in AUD pilot grant (U54 AA027989). We want to acknowledge the participants in the Yale-Penn cohorts. The authors are supported by grants from the National Institutes of Health (NIH) (R01 AA026364, R01 DA037974, P50 AA012870, R21 CA252916, U54 AA027989R01AA026364, R01AA11330, R01DA12890, R01DA037974, P50AA012870, R21CA252916, U54AA027989, RM1 HG011558, and R01 AA030056) and the Department of Veterans Affairs (1I01CX001849 and I01 BX004820). H.Z. was also supported by a NARSAD Young Investigator grant #27835 from the Brain & Behavior Research Foundation.

## References

1. Collaborators GBDA. Population-level risks of alcohol consumption by amount, geography, age, sex, and year: a systematic analysis for the Global Burden of Disease Study 2020. Lancet. 2022;400(10347):185–235.

2. Verhulst B, Neale MC, Kendler KS. The heritability of alcohol use disorders: a meta-analysis of twin and adoption studies. Psychol Med. 2015;45(5):1061–1072.

3. Harada S, Agarwal DP, Goedde HW, Tagaki S, Ishikawa B. Possible protective role against alcoholism for aldehyde dehydrogenase isozyme deficiency in Japan. Lancet. 1982;2(8302):827.

4. Borras E, Coutelle C, Rosell A, et al. Genetic polymorphism of alcohol dehydrogenase in europeans: the ADH2*2 allele decreases the risk for alcoholism and is associated with ADH3*1. Hepatology. 2000;31(4):984–989.

5. Li D, Zhao H, Gelernter J. Strong protective effect of the aldehyde dehydrogenase gene (ALDH2) 504lys (*2) allele against alcoholism and alcohol-induced medical diseases in Asians. Hum Genet. 2012;131(5):725–737.

6. Li D, Zhao H, Gelernter J. Strong association of the alcohol dehydrogenase 1B gene (ADH1B) with alcohol dependence and alcohol-induced medical diseases. Biol Psychiatry. 2011;70(6):504–512.

7. Edenberg HJ. The genetics of alcohol metabolism: role of alcohol dehydrogenase and aldehyde dehydrogenase variants. Alcohol Res Health. 2007;30(1):5–13.

8. Gelernter J, Polimanti R. Genetics of substance use disorders in the era of big data. Nat Rev Genet. 2021;22(11):712–729.

9. Frank J, Cichon S, Treutlein J, et al. Genome-wide significant association between alcohol dependence and a variant in the ADH gene cluster. Addict Biol. 2012;17(1):171–180.

10. Park BL, Kim JW, Cheong HS, et al. Extended genetic effects of ADH cluster genes on the risk of alcohol dependence: from GWAS to replication. Hum Genet. 2013;132(6):657–668.

11. Gelernter J, Kranzler HR, Sherva R, et al. Genome-wide association study of alcohol dependence:significant findings in African- and European-Americans including novel risk loci. Mol Psychiatry. 2014;19(1):41–49.

12. Walters RK, Polimanti R, Johnson EC, et al. Transancestral GWAS of alcohol dependence reveals common genetic underpinnings with psychiatric disorders. Nat Neurosci. 2018;21(12):1656–1669.

13. Sanchez-Roige S, Palmer AA, Fontanillas P, et al. Genome-Wide Association Study Meta-Analysis of the Alcohol Use Disorders Identification Test (AUDIT) in Two Population-Based Cohorts. Am J Psychiatry. 2019;176(2):107–118.

14. Kranzler HR, Zhou H, Kember RL, et al. Genome-wide association study of alcohol consumption and use disorder in 274,424 individuals from multiple populations. Nat Commun. 2019;10(1):1499.

15. Zhou H, Sealock JM, Sanchez-Roige S, et al. Genome-wide meta-analysis of problematic alcohol use in 435,563 individuals yields insights into biology and relationships with other traits. Nat Neurosci. 2020;23(7):809–818.

16. Koob GF, Volkow ND. Neurocircuitry of addiction. Neuropsychopharmacology. 2010;35(1):217–238.

17. Zhou H, Kember RL, Deak JD, et al. Multi-ancestry study of the genetics of problematic alcohol use in over 1 million individuals. Nat Med. 2023;29(12):3184–3192.

18. Palmer DS, Howrigan DP, Chapman SB, et al. Exome sequencing in bipolar disorder identifies AKAP11 as a risk gene shared with schizophrenia. Nature genetics. 2022;54(5):541–547.

19. Jurgens SJ, Choi SH, Morrill VN, et al. Analysis of rare genetic variation underlying cardiometabolic diseases and traits among 200,000 individuals in the UK Biobank. Nature genetics. 2022;54(3):240–250.

20. Wang Q, Dhindsa RS, Carss K, et al. Rare variant contribution to human disease in 281,104 UK Biobank exomes. Nature. 2021;597(7877):527-532.

21. Backman JD, Li AH, Marcketta A, et al. Exome sequencing and analysis of 454,787 UK Biobank participants. Nature. 2021;599(7886):628-634.

22. Singh T, Poterba T, Curtis D, et al. Rare coding variants in ten genes confer substantial risk for schizophrenia. Nature. 2022;604(7906):509-516.

23. Wainschtein P, Jain D, Zheng Z, et al. Assessing the contribution of rare variants to complex trait heritability from whole-genome sequence data. Nat Genet. 2022;54(3):263–273.

24. Jin SC, Homsy J, Zaidi S, et al. Contribution of rare inherited and de novo variants in 2,871 congenital heart disease probands. Nature genetics. 2017;49(11):1593–1601.

25. Jang SK, Evans L, Fialkowski A, et al. Rare genetic variants explain missing heritability in smoking. Nat Hum Behav. 2022;6(11):1577–1586.

26. Gentry AE, Alexander JC, Ahangari M, et al. Case-only exome variation analysis of severe alcohol dependence using a multivariate hierarchical gene clustering approach. PLoS One. 2023;18(4):e0283985.

27. Hill SY, Hostyk J. A whole exome sequencing study to identify rare variants in multiplex families with alcohol use disorder. Front Psychiatry. 2023;14:1216493.

28. Bycroft C, Freeman C, Petkova D, et al. The UK Biobank resource with deep phenotyping and genomic data. Nature. 2018;562(7726):203-209.

29. American Psychiatric Association. Diagnostic and Statistical Manual of Mental Disorders 4th Ed. American Psychiatric Press: Washington, DC, USA. 1994.

30. Pierucci-Lagha A, Gelernter J, Feinn R, et al. Diagnostic reliability of the Semi-structured Assessment for Drug Dependence and Alcoholism (SSADDA). Drug Alcohol Depend. 2005;80(3):303–312.

31. Van der Auwera GA, Carneiro MO, Hartl C, et al. From FastQ data to high confidence variant calls: the Genome Analysis Toolkit best practices pipeline. Curr Protoc Bioinformatics. 2013;43(1110):11 10 11-11 10 33.

32. Lek M, Karczewski KJ, Minikel EV, et al. Analysis of protein-coding genetic variation in 60,706 humans. Nature. 2016;536(7616):285-291.

33. Chen S, Francioli LC, Goodrich JK, et al. A genomic mutational constraint map using variation in 76,156 human genomes. Nature. 2024;625(7993):92-100.

34. Genomes Project C, Auton A, Brooks LD, et al. A global reference for human genetic variation. Nature. 2015;526(7571):68-74.

35. Wang K, Li M, Hakonarson H. ANNOVAR: functional annotation of genetic variants from high-throughput sequencing data. Nucleic acids research. 2010;38(16):e164.

36. Ioannidis NM, Rothstein JH, Pejaver V, et al. REVEL: An Ensemble Method for Predicting the Pathogenicity of Rare Missense Variants. Am J Hum Genet. 2016;99(4):877–885.

37. Ghosh R, Oak N, Plon SE. Evaluation of in silico algorithms for use with ACMG/AMP clinical variant interpretation guidelines. Genome Biol. 2017;18(1):225.

38. Ma C, Blackwell T, Boehnke M, Scott LJ, Go TDi. Recommended joint and meta-analysis strategies for case-control association testing of single low-count variants. Genet Epidemiol. 2013;37(6):539–550.

39. Zhou W, Bi W, Zhao Z, et al. SAIGE-GENE+ improves the efficiency and accuracy of set-based rare variant association tests. Nature genetics. 2022;54(10):1466–1469.

40. Karczewski KJ, Solomonson M, Chao KR, et al. Systematic single-variant and gene-based association testing of thousands of phenotypes in 394,841 UK Biobank exomes. Cell Genom. 2022;2(9):100168.

41. Ng SB, Buckingham KJ, Lee C, et al. Exome sequencing identifies the cause of a mendelian disorder. Nat Genet. 2010;42(1):30–35.

42. Dutta N, Helton SG, Schwandt M, Zhu X, Momenan R, Lohoff FW. Genetic Variation in the Vesicular Monoamine Transporter 1 (VMAT1/SLC18A1) Gene and Alcohol Withdrawal Severity. Alcohol Clin Exp Res. 2016;40(3):474–481.

43. Stankiewicz TR, Linseman DA. Rho family GTPases: key players in neuronal development, neuronal survival, and neurodegeneration. Front Cell Neurosci. 2014;8:314.

44. Genetic Modifiers of Huntington’s Disease Consortium. Electronic address ghmhe, Genetic Modifiers of Huntington’s Disease C. CAG Repeat Not Polyglutamine Length Determines Timing of Huntington’s Disease Onset. Cell. 2019;178(4):887-900 e814.

45. Zheng C, Zheng Z, Zhang Z, et al. IFIT5 positively regulates NF-kappaB signaling through synergizing the recruitment of IkappaB kinase (IKK) to TGF-beta-activated kinase 1 (TAK1). Cell Signal. 2015;27(12):2343–2354.

46. Lo UG, Pong RC, Yang D, et al. IFNgamma-Induced IFIT5 Promotes Epithelial-to-Mesenchymal Transition in Prostate Cancer via miRNA Processing. Cancer Res. 2019;79(6):1098–1112.

47. Dai Q, Pu SS, Yang X, et al. Whole Transcriptome Sequencing of Peripheral Blood Shows That Immunity/GnRH/PI3K-Akt Pathways Are Associated With Opioid Use Disorder. Front Psychiatry. 2022;13:893303.

48. Ray PR, Shiers S, Caruso JP, et al. RNA profiling of human dorsal root ganglia reveals sex differences in mechanisms promoting neuropathic pain. Brain. 2023;146(2):749–766.

49. Zapata RC, Nasamran CA, Chilin-Fuentes DR, Dulawa SC, Osborn O. Identification of adipose tissue transcriptomic memory of anorexia nervosa. Mol Med. 2023;29(1):109.

50. Solmi M, Wade TD, Byrne S, et al. Comparative efficacy and acceptability of psychological interventions for the treatment of adult outpatients with anorexia nervosa: a systematic review and network meta-analysis. Lancet Psychiatry. 2021;8(3):215–224.

51. Zhang Z, Yang H, Wang H. The histone H2A deubiquitinase USP16 interacts with HERC2 and fine-tunes cellular response to DNA damage. J Biol Chem. 2014;289(47):32883–32894.

52. Tian C, Duan L, Fu C, He J, Dai J, Zhu G. Study on the Correlation Between Iris Characteristics and Schizophrenia. Neuropsychiatr Dis Treat. 2022;18:811–820.

53. Yang L, Zhan GD, Ding JJ, et al. Psychiatric illness and intellectual disability in the Prader-Willi syndrome with different molecular defects--a meta analysis. PLoS One. 2013;8(8):e72640.

54. Sinnema M, Boer H, Collin P, et al. Psychiatric illness in a cohort of adults with Prader-Willi syndrome. Res Dev Disabil. 2011;32(5):1729–1735.

55. Li H, Durbin R. Fast and accurate short read alignment with Burrows-Wheeler transform. Bioinformatics. 2009;25(14):1754–1760.

56. Dao C, Zhou H, Small A, et al. The impact of removing former drinkers from genome-wide association studies of AUDIT-C. Addiction. 2021;116(11):3044–3054.

57. Willer CJ, Li Y, Abecasis GR. METAL: fast and efficient meta-analysis of genomewide association scans. Bioinformatics. 2010;26(17):2190–2191.

