## Supplementary Figures 1-7 for "Multi-ancestry Whole-exome Sequencing Study of Alcohol Use Disorder in Two Cohorts"

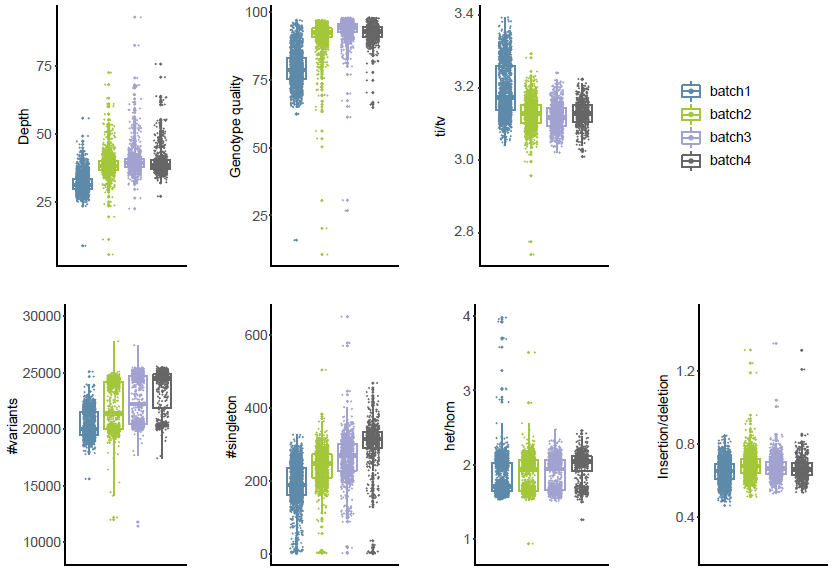

**Supplementary Figure 1. Distributions of the sequencing quality metrics of YP WES data.** Het/Hom, ratio of the heterozygous variants to nonreference homozygous variants. Ti/Tv, the ratio of transitions to transversions.

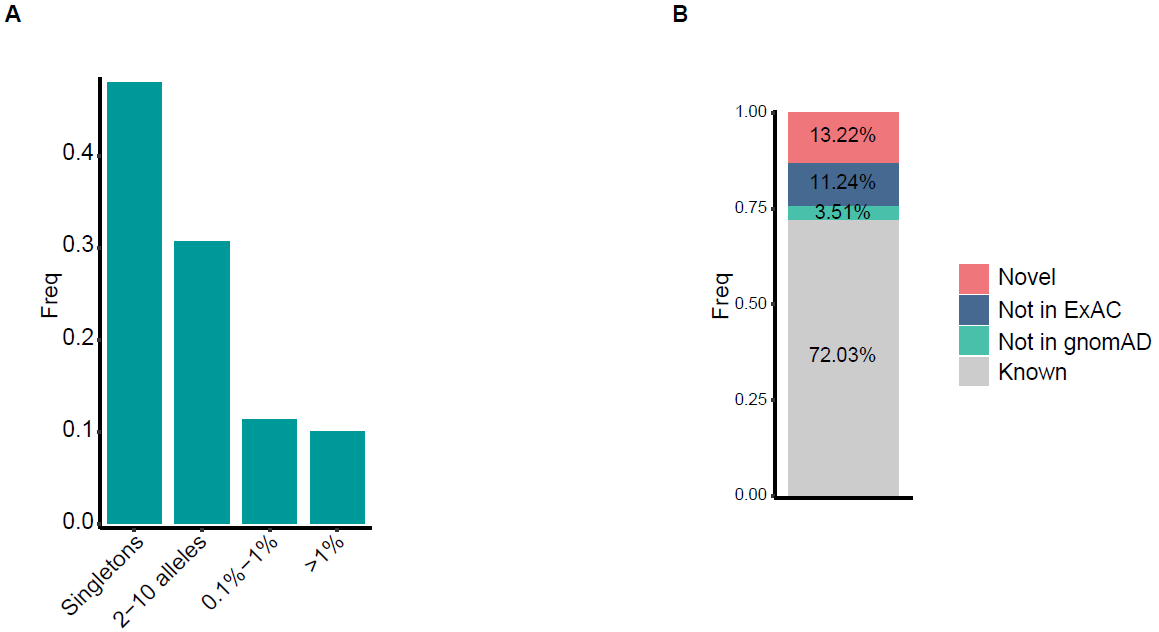

**Supplementary Figure 2. The allele frequency spectrum of variants and novel variants identified in the YP dataset.** Proportions of variants from YP that are not identified by either ExAC or whole-genome sequences from gnomAD v3.1.2, exist only in one of them or both were indicated.

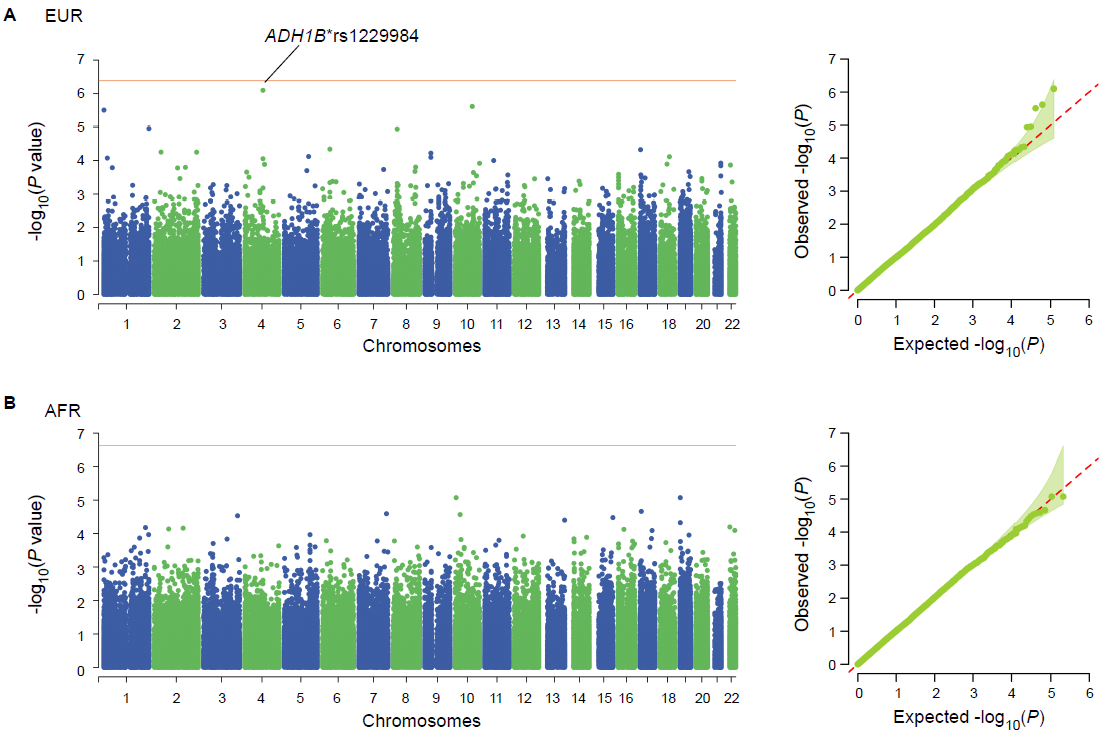

**Supplementary Figure 3. Single-variant association analysis results of the YP dataset.** Manhattan plot and QQ plot for EUR (A) and AFR (B) in YP.

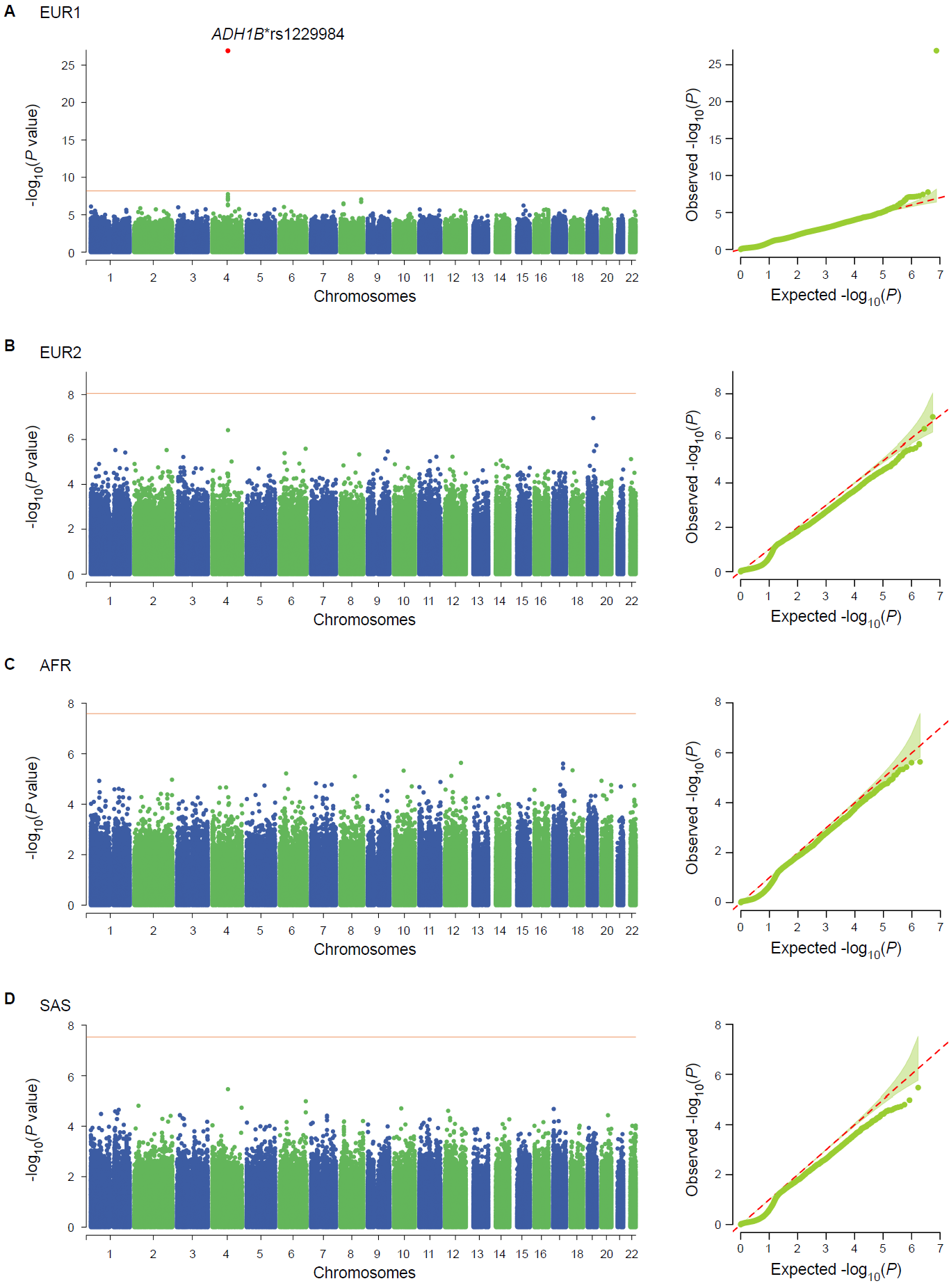

**Supplementary Figure 4. Single-variant association analysis in UKB.** Manhattan plot and QQ plot for EUR1 subpopulation, EUR2 subpopulation, AFR subpopulation and SAS subpopulation from upper panel to lower panel.

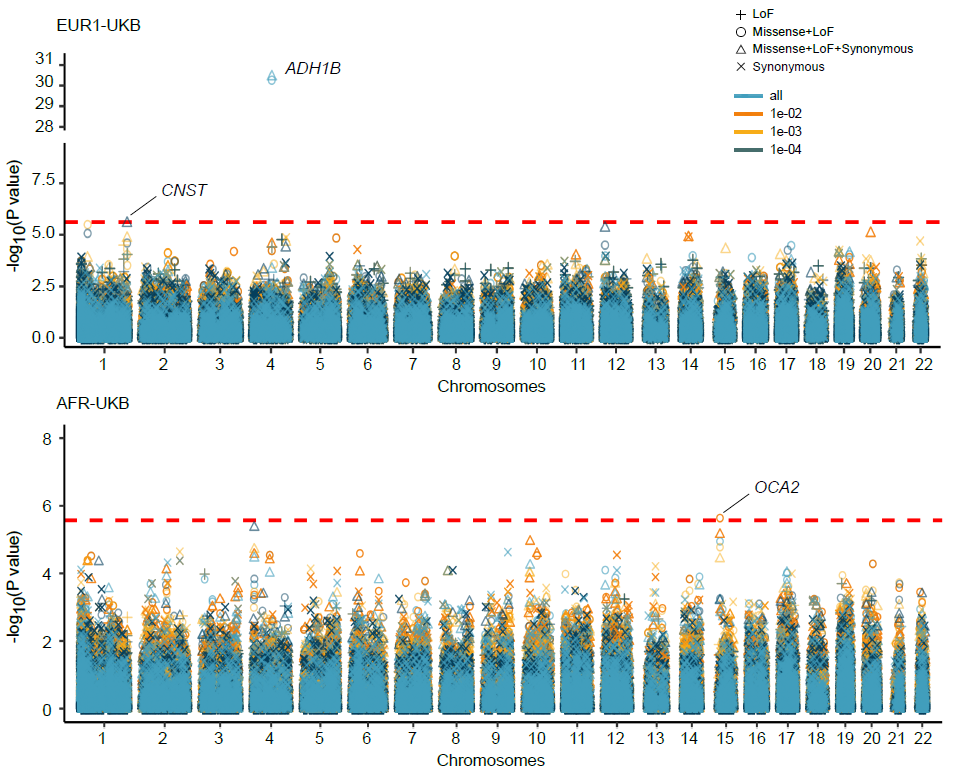

**Supplementary Figure 5. Gene-based association analysis for EUR1 and AFR subpopulation in UKB.** Manhattan plots showed the combined gene-based results under each MAF and variant class combination groups. Red dashed horizontal line represents exome-wide significance under Bonferroni correction. Signals passed the threshold were highlighted in red. A signal from *CNST* gene almost reached the Bonferroni-corrected exome-wide significance in EUR1 subpopulation was marked. Manhattan plots were not presented for EUR2 and SAS due to no significant results from these two subpopulations after Bonferroni corrections.

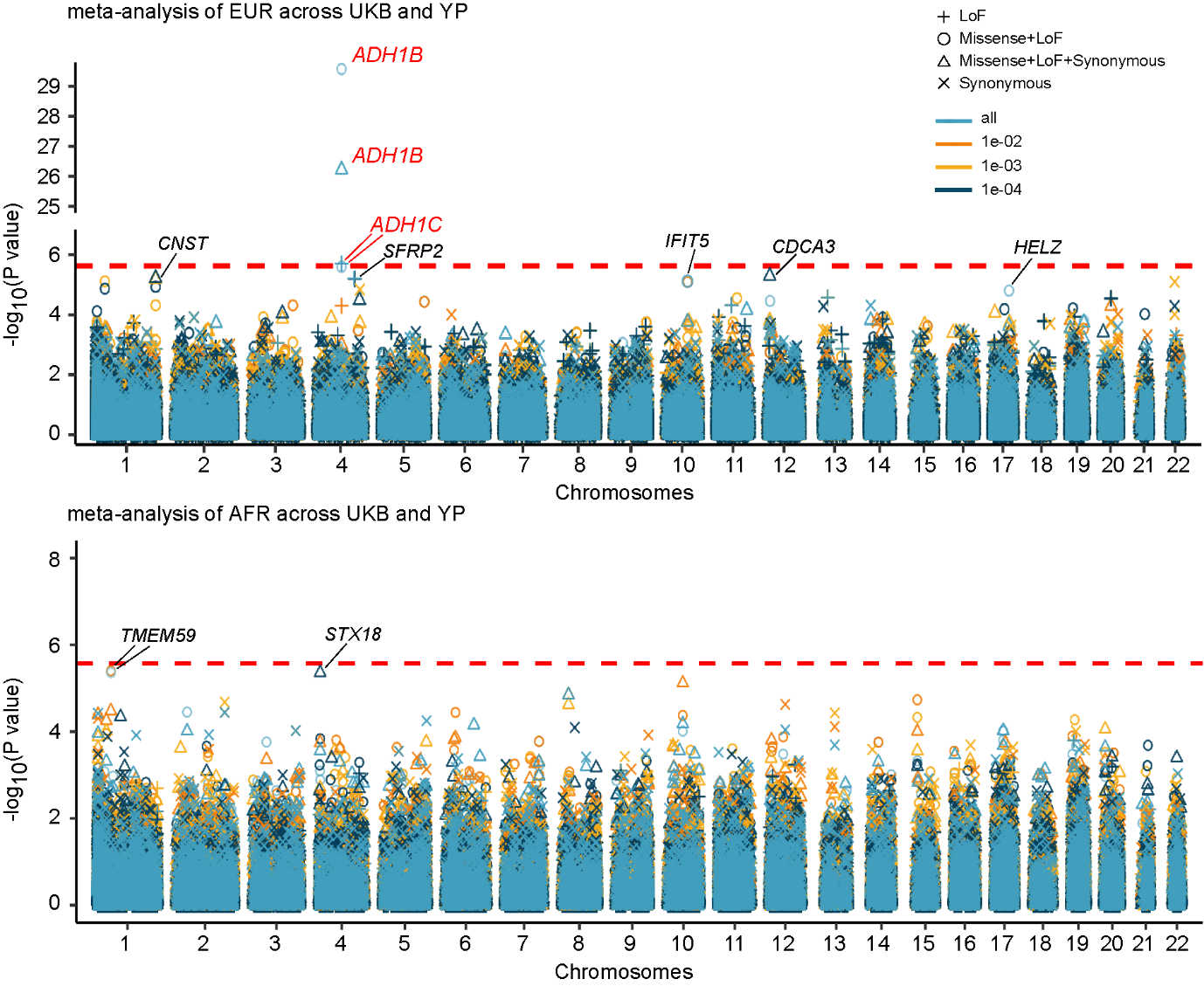

**Supplementary Figure 6. Within-ancestry meta-analyses of the gene-based association results for EUR and AFR across YP and UKB.** Manhattan plots showed the combined gene-based results under each MAF and variant class combination groups. Red dashed horizontal line represents exome-wide significance under Bonferroni correction. Markers passed the Bonferroni-corrected threshold were highlighted in red. Markers passed FDR-corrected *P*-value<0.1 were marked in black. MAF groups of cutoff 0.0001 were not considered in the meta-analyses because there were no variants under the MAF cutoff of 0.0001 in YP cohort.

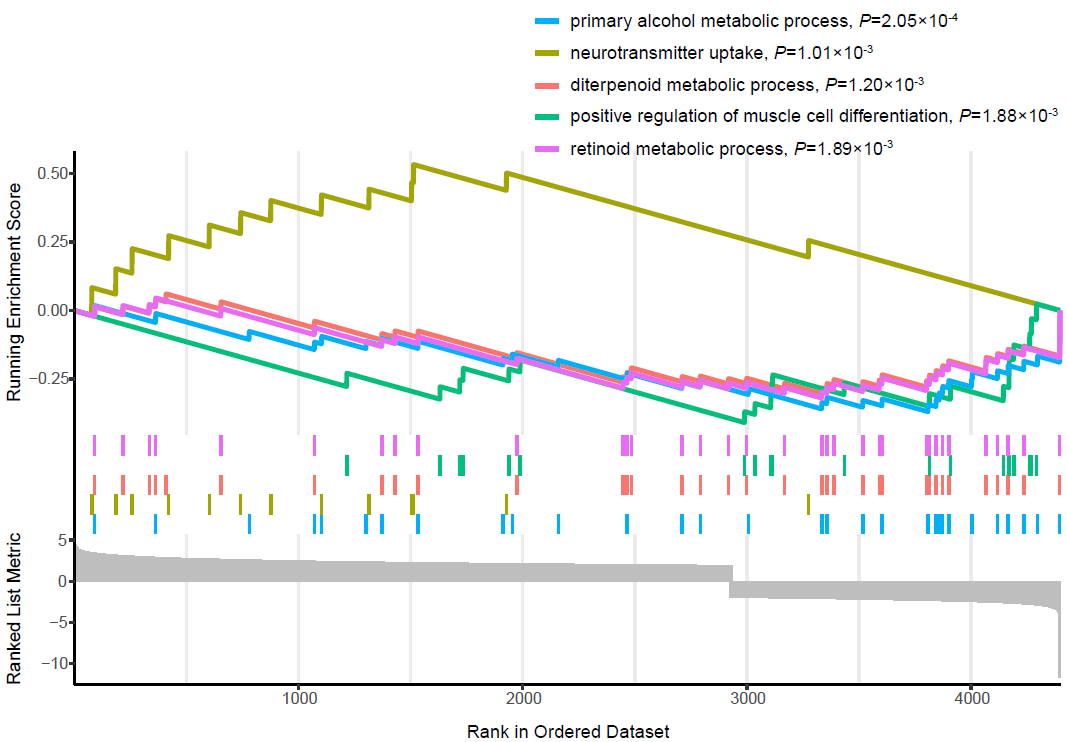

**Supplementary Figure 7. GO enrichment analysis for nominally significant genes from the cross-ancestry meta-analysis of the gene-based association results.** Genes that were nominally significant in at least one of the MAF and variant class combination groups were selected. Selected genes were ranked based on the meta-analysis Z-scores. Each vertical line in the figure represents one gene. The top 5 nominally significant GO terms were shown.
